# Urban Sprawl of Covid-19 Epidemic in India: Lessons in the First Semester

**DOI:** 10.1101/2020.08.17.20176537

**Authors:** Rajeev Gupta, Kiran Gaur, Raghubir S Khedar, Rajinder K Dhamija

**Affiliations:** Department of Medicine, Eternal Heart Care Centre & Research Institute, Jaipur, India; Department of Statistics, Mathematics and Computer Science, SKN College of Agriculture, SKN Agriculture University, Jobner, Jaipur, India.; Department of Neurology, Lady Hardinge Medical College & SSK Hospital, New Delhi, India.

**Keywords:** Covid-19, SARS, Coronavirus, India, Epidemiology, Urban Health

## Abstract

**Background & Objective:** The covid-19 epidemic is rapidly escalating in India and unlike developed countries there is no evidence of plateau or decline in the past 6 months. To evaluate association of state-level sociodemographics with incident cases and deaths we performed an ecological study.

**Methods:** Publicly available data sources were used. Absolute number of covid-19 cases and deaths were obtained and cases and deaths/million in each state calculated from February to July 2020. To assess association of state level disease burden with sociodemographic variables (urbanization, human development, healthcare availability, healthcare access and quality etc.) we determined Pearson’s correlation and logarithmic trends.

**Results:** Covid-19 in India has led to >2,000,000 cases and 45,000 deaths by end July 2020. There is large variation in state-level cases/million ranging from 7247 (Delhi), 3728 (Goa) and 3427 (Maharashtra) to less than 300/million in a few. Deaths/million range from 212 (Delhi), 122 (Maharashtra) and 51 (Tamilnadu) to 2 in north-eastern states. Most of the high burden states (except Delhi) are reporting increasing burden and deaths with the largest increase in July 2020. There is a significant positive correlation of urbanization with covid-19 cases (r=0.65, R^2^=0.35) and deaths (r=0.60, R^2^=0.28) and weaker correlation with other sociodemographic variables. From March to July 2020, stable R^2^ value for urbanization is observed with cases (0.37 to 0.39) while it is increasing for deaths (0.10 to 0.28).

**Conclusions:** Covid-19 epidemic is escalating in India and cases as well as deaths are significantly greater in more urbanized states. Prevention, control and treatment should focus on urban health systems.

## INTRODUCTION

Severe acute respiratory syndrome (SARS) related to novel coronavirus (covid-19) has rapidly spread globally. Initial cases were reported from China from where it rapidly spread to Europe, East Asia, Europe, North and South America and lately to Africa and South Asia.^1^ India is currently facing the maximum impact.^2,3^ The disease has been an urban phenomenon globally and has been mainly reported from densely populated urban conglomerations in China and low socioeconomic status urban locations in Europe and North America.^4^ United Nations estimates that 90% of all Covid-19 cases are urban and there is only limited evidence of rural spread.^5^ Covid-19 related burden has been controlled in many countries across the globe using well known public health strategies of testing and tracing of cases (using laboratory methods) and isolation.^6^ This strategy combined with universal masking, hand hygiene and sanitation, avoidance of crowding and physical distancing has led to virtual disappearance of covid-19 from some countries.^7^ Drug therapies for prophylaxis or treatment are yet not available as none has proved successful in randomised controlled trials.^8^ Vaccine is under development.

In India, the epidemic is now a semester (6 months) old. Epidemiological transition of the disease from contacts of returning travellers from Asia and Europe to metropolitan cities and large townships has been reported.^9^ The disease has now transited from the major metropolises to smaller cities, towns and townships with limited spread to the rural.^10^ The disease initially presented in low socioeconomic urban locations and gradually spread to rural locations due to large scale urban to rural migration.^9^ However, it remained ensconced in urban areas also and with the release of government-instituted lockdown and subsequent crowding the epidemic has resurfaced in urban locations.^11^ This is unlike the olden epidemics whence disease would transmit from the rich to poor people and urban to rural locations.^12,13^ Understanding macrolevel determinants of this transition is important for understanding covid-19 dynamics and to formulate policies for controlling it. In the present article we describe covid-19 related disease and death burden in terms of absolute numbers and per million populations in all states of the country using publicly available data over the first semester of the India epidemic, February to July 2020. We have also estimated association of disease burden and mortality with various sociodemographic indices using univariate analysis to identify important drivers.

## METHODS

The study has been conducted using publicly available data.^9,14^ The project proposal was submitted to institutional ethics committee as part of covid-19 registry being maintained at our centre. The committee approved use of secondary data for this report. Daily data on covid-19 in various states and regions of India are being regularly updated at a non-commercial public website, https://www.covid19india.org.^14^ This website updates daily data on covid-19 related cases, deaths, recovery and testing at the state level of India. We obtained data for all the states in the country and clubbed daily data into weekly and monthly numbers beginning February 2020 to end of July 2020. The data were collated on spreadsheets. We then calculated number of cases and deaths per million population (2020 estimates) for each state. We also obtained data on multiple sociodemographic indices of each state from public websites. The following indices were used: urbanization index (UI, proportion of urban to rural population), human development index (HDI), social development index (SDI),sociodemographic index (SI), epidemiological transition index (ETI, proportion of disability adjusted life years due to communicable, maternal, neonatal and nutritional diseases to non-communicable diseases), healthcare availability and quality index (HAQI), healthcare availability index (HAI), and social vulnerability index(VI). (Supplementary Table 1). Details of estimation of each of these indices have been reported earlier.^15,16^

To determine association of the state-level sociodemographic variables with covid-19 cases and deaths we initially calculated Pearson’s correlation coefficient (r value) using MS Office Excel-2007. MS Office Powerpoint-2007 was used to plot scatter-graphs for estimation of correlation of various sociodemographic indices with total cases/million and deaths/million at end of 6 months of the data (July 2020). Logarithmic trend-line was drawn to calculate trends (R^2^). SPSS Statistical Package was used to calculate univariate and multivariate regression association statistics. To identify monthly change in sociodemographic association with covid-19 cases/million and deaths/million we calculated R^2^ values for months of April, May, June and July.

## RESULTS

India is currently (July 2020) reporting more than 50,000 covid-9 cases and 1,000 covid-19 related deaths daily and the cumulative burden is high for cases (>1,800,000) as well as deaths (38,000). India is among the top-5 countries in terms of absolute number of cases and deaths.^17^ Although, the number of cases and deaths per million (cases 1300, deaths 30) is low, it is the only large country where the epidemic is escalating.

There is significant state-level variation with five states- Maharashtra, Tamilnadu, Andhra Pradesh, Delhi and Karnataka accounting for more than two-thirds of cases and deaths (Figure 1). Cases and deaths per million also show large variation with cases per million highest in Delhi (7247), Goa (3728), Maharashtra (3427), Tamilnadu (3158) and Andhra Pradesh (2614) and deaths per million the highest in Delhi (212), Maharashtra (122), Tamilnadu (51), Gujarat (38) and Pondicherry (35) (Figure 2). Absolute number and numbers/million population of monthly covid-19 cases in various states of the country are shown in Table 1.The cases were low in Feb to May 2020 but have increased rapidly in the following months with greatest escalation in July 2020. Significant diversity is observed in trends of increase in monthly number of cases and cases/million across various states of the country. Greatest month-on-month increase is observed in a few states: Delhi, Goa, Maharashtra, Tamilnadu and Andhra Pradesh (Figure 3). Absolute number of deaths/month and death/million/month are shown in Table 2. Similar to cases, deaths are the highest in Maharashtra, Delhi, Tamilnadu, Gujarat and Karnataka while deaths/million are the highest in Delhi, Maharashtra, Tamilnadu, Gujarat and Pondicherry. State-level trends in monthly deaths/million reveal rapidly escalating trends in Delhi, Maharashtra, Tamilnadu, Gujarat and Pondicherry. ON the other hand, a decline in trends of deaths/million is observed in Delhi and Andhra Pradesh (Figure 3).

**Figure 1:**
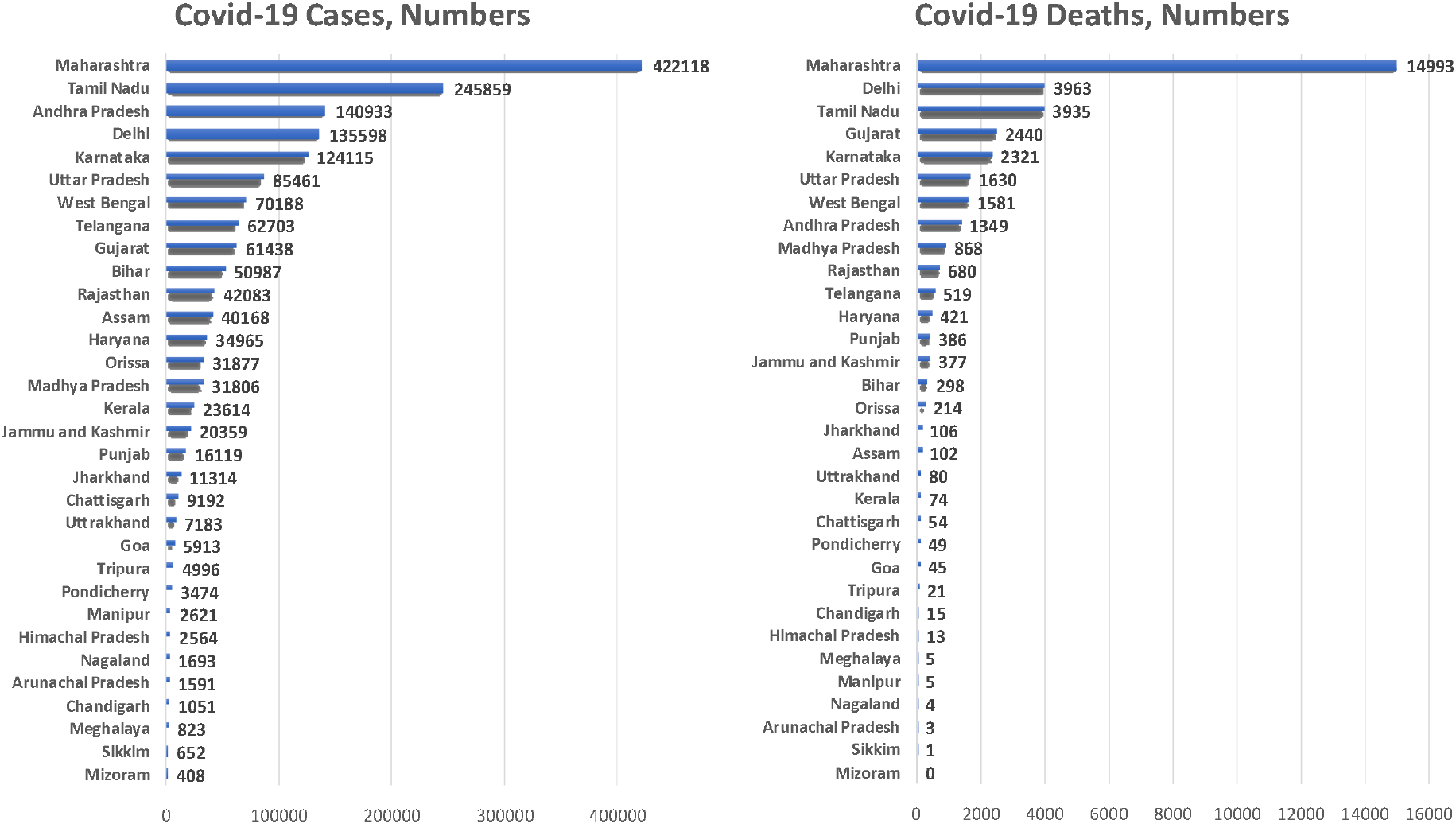
Absolute number of cumulative covid-19 cases and deaths in various states (July 2020)

**Figure 2:**
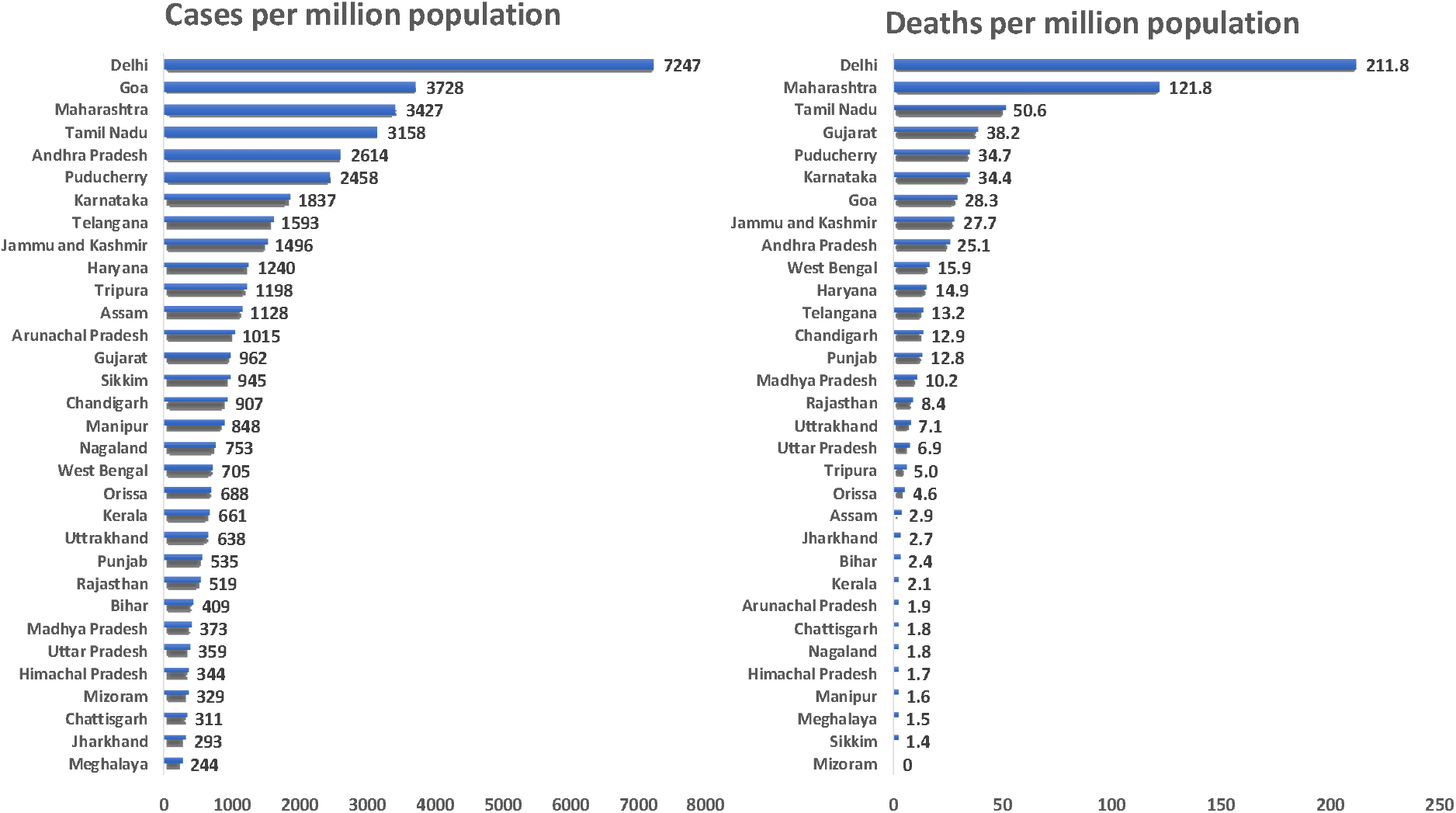
Covid-19 cumulative cases and deaths per million in various states (July 2020)

**Table 1:**
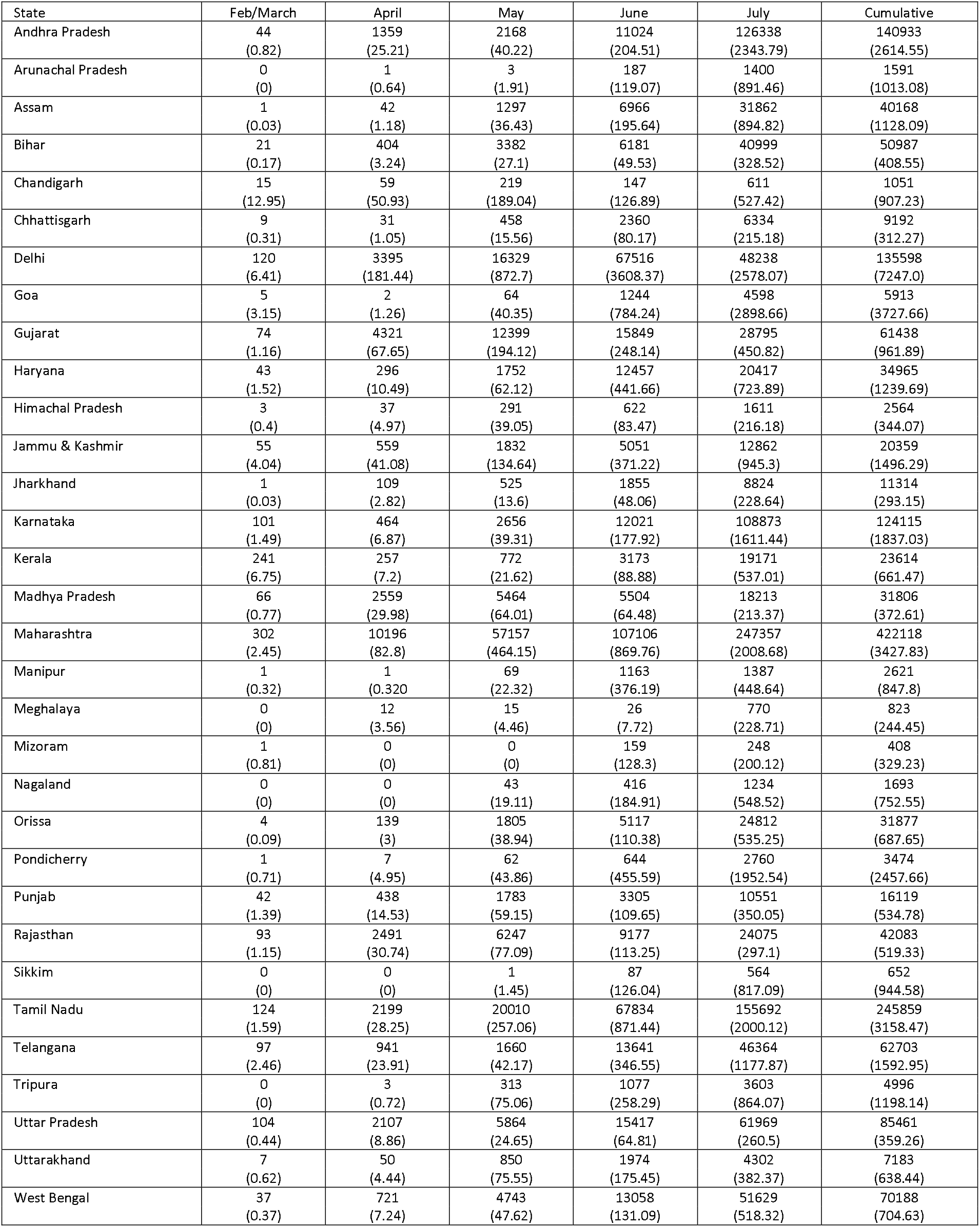
Monthly covid-19 cases in absolute numbers and cases/million in various states.

**Figure 3:**
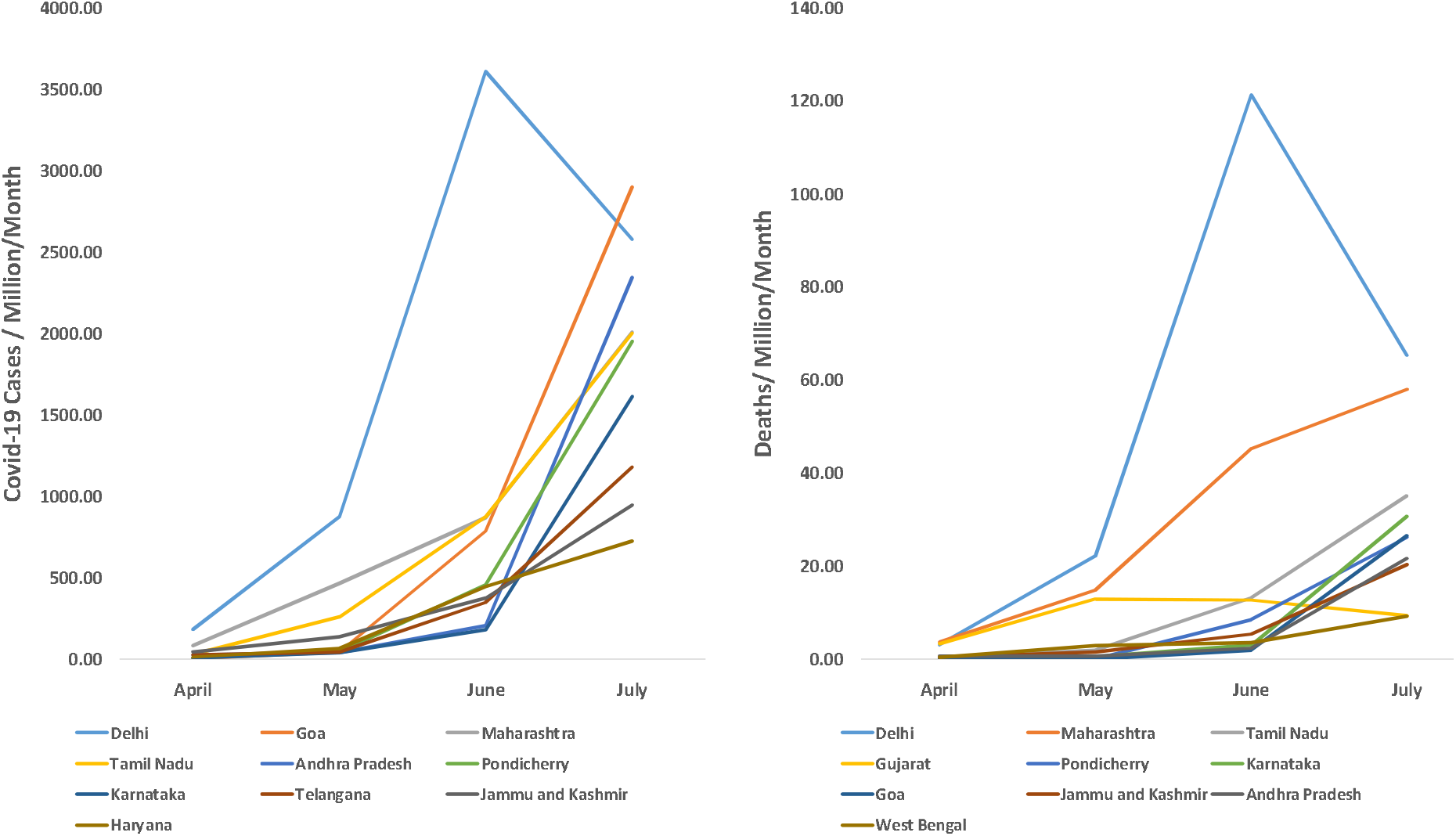
Trends in monthly cases/million and deaths/million in rapidly increasing states from Feb/March to July 2020 illion.

**Table 2:** Monthly Covid-19 deaths, absolute numbers and (deaths/million) in various states of India.

| State | Feb/March | April | May | June | July | Cumulative |
| --- | --- | --- | --- | --- | --- | --- |
| Andhra Pradesh | 0<br>(0) | 31<br>(0.58) | 31<br>(0.58) | 125<br>(2.32) | 1162<br>(21.56) | 1349<br>(25.03) |
| Arunachal Pradesh | 0<br>(0) | 0<br>(0) | 0<br>(0) | 1<br>(0.64) | 2<br>(1.27) | 3<br>(1.91) |
| Assam | 0<br>(0) | 1<br>(0.03) | 2<br>(0.06) | 8<br>(0.22) | 91<br>(2.56) | 102<br>(2.86) |
| Bihar | 1<br>(0.01) | 1<br>(0.01) | 21<br>(0.17) | 45<br>(0.36) | 230<br>(1.84) | 298<br>(2.39) |
| Chandigarh | 0<br>(0) | 0<br>(0) | 4<br>(3.45) | 2<br>(1.73) | 9<br>(7.77) | 15<br>(12.95) |
| Chattisgarh | 0<br>(0) | 0<br>(0) | 1<br>(0.03) | 12<br>(0.41) | 41<br>(1.39) | 54<br>(1.83) |
| Delhi | 2<br>(0.11) | 57<br>(3.05) | 414<br>(22.13) | 2269<br>(121.27) | 1221<br>(65.26) | 3963<br>(211.8) |
| Goa | 0<br>(0) | 0<br>(0) | 0<br>(0) | 3<br>(1.89) | 42<br>(26.48) | 45<br>(28.37) |
| Gujarat | 6<br>(0.09) | 208<br>(3.26) | 824<br>(12.9) | 810<br>(12.68) | 592<br>(9.27) | 2440<br>(38.2) |
| Haryana | 0<br>(0) | 4<br>(0.14) | 16<br>(0.57) | 216<br>(7.66) | 185<br>(6.56) | 421<br>(14.93) |
| Himachal Pradesh | 1<br>(0.13) | 1<br>(0.13) | 4<br>(0.54) | 3<br>(0.4) | 4<br>(0.54) | 13<br>(1.74) |
| Jammu & Kashmir | 2<br>(0.15) | 6<br>(0.44) | 20<br>(1.47) | 73<br>(5.37) | 276<br>(20.28) | 377<br>(27.71) |
| Jharkhand | 0<br>(0) | 3<br>(0.08) | 2<br>(0.05) | 10<br>(0.26) | 91<br>(2.36) | 106<br>(2.75) |
| Karnataka | 3<br>(0.04) | 19<br>(0.28) | 29<br>(0.43) | 197<br>(2.92) | 2073<br>(30.68) | 2321<br>(34.35) |
| Kerala | 2<br>(0.06) | 2<br>(0.06) | 6<br>(0.17) | 15<br>(0.42) | 49<br>(1.37) | 74<br>(2.07) |
| Madhya Pradesh | 5<br>(0.06) | 133<br>(1.56) | 213<br>(2.5) | 222<br>(2.6) | 295<br>(3.46) | 868<br>(10.17) |
| Maharashtra | 10<br>(0.08) | 448<br>(3.64) | 1827<br>(14.84) | 5569<br>(45.22) | 7139<br>(57.97) | 14993<br>(121.75) |
| Manipur | 0<br>(0) | 0<br>(0) | 0<br>(0) | 0<br>(0) | 5<br>(1.62) | 5<br>(1.62) |
| Meghalaya | 0<br>(0) | 1<br>(0.3) | 0<br>(0) | 0<br>(0) | 4<br>(1.19) | 5<br>(1.49) |
| Mizoram | 0<br>(0) | 0<br>(0) | 0<br>(0) | 0<br>(0) | 0<br>(0) | 0<br>(0) |
| Nagaland | 0<br>(0) | 0<br>(0) | 0<br>(0) | 0<br>(0) | 4<br>(1.78) | 4<br>(1.78) |
| Orissa | 0<br>(0) | 1<br>(0.02) | 8<br>(0.17) | 23<br>(0.5) | 182<br>(3.93) | 214<br>(4.62) |
| Pondicherry | 0<br>(0) | 0<br>(0) | 0<br>(0) | 12<br>(8.49) | 37<br>(26.18) | 49<br>(34.66) |
| Punjab | 4<br>(0.13) | 16<br>(0.53) | 25<br>(0.83) | 99<br>(3.28) | 242<br>(8.03) | 386<br>(12.81) |
| Rajasthan | 0<br>(0) | 58<br>(0.72) | 136<br>(1.68) | 219<br>(2.7) | 267<br>(3.29) | 680<br>(8.39) |
| Sikkim | 0<br>(0) | 0<br>(0) | 0<br>(0) | 0<br>(0) | 1<br>(1.45) | 1<br>(1.45) |
| Tamil Nadu | 1<br>(0.01) | 26<br>(0.33) | 149<br>(1.91) | 1025<br>(13.17) | 2734<br>(35.12) | 3935<br>(50.55) |
| Telangana | 6<br>(0.15) | 22<br>(0.56) | 54<br>(1.37) | 178<br>(4.52) | 259<br>(6.58) | 519<br>(13.19) |
| Tripura | 0<br>(0) | 0<br>(0) | 0<br>(0) | 1<br>(0.24) | 20<br>(4.8) | 21<br>(5.04) |
| Uttar Pradesh | 0<br>(0) | 40<br>(0.17) | 177<br>(0.74) | 480<br>(2.02) | 933<br>(3.92) | 1630<br>(6.85) |
| Uttarakhand | 0<br>(0) | 0<br>(0) | 5<br>(0.44) | 36<br>(3.2) | 39<br>(3.47) | 80<br>(7.11) |
| West Bengal | 3<br>(0.03) | 30<br>(0.3) | 284<br>(2.85) | 351<br>(3.52) | 913<br>(9.17) | 1581<br>(15.87) |

We correlated cumulative covid-19 cases and deaths/million at July 2020 with various sociodemographic indices (Table 3). A significant correlation of state-level urbanization index is observed with cases/million (r = 0.58, R^2^ = 0.35) as well as deaths/million (r = 0.55, R^2^ = 0.28) (Figure 4). Significant correlation (r value) is also observed for cases/million with HDI (0.42), sociodemographic index (0.60), ETI (−0.44) and HAQI (0.50) and non-significant correlation with SDI (−0.20), HAI (0.12), and VI (0.01). Significant correlation for deaths/million is observed for UI and sociodemographic index (0.50) only (Table 3). Multiple regression analysis reveals that the only significant association for covid-19 related deaths is with urbanization index (B = 0.55+0.24, standardized beta = 0.52, p = 0.040).

**Table 3:** Correlation of state-level covid-19 cases/million and deaths/million with various sociodemographic indices (Pearson’s correlation) in July 2020.

|  | Source | Cases/million | Deaths/million |
| --- | --- | --- | --- |
| Urbanization index (UI) | Census of India | 0.65** | 0.60** |
| Human development index (HDI) | Government of India | 0.42** | 0.32 |
| Social development index (SDI) | Government of India | -0.20 | -0.13 |
| Sociodemographic index (SI) | Global Burden of Disease study | 0.60** | 0.50** |
| Epidemiological transition index (ETI) | Global Burden of Disease Study | -0.44** | -0.30 |
| Healthcare availability index (HAI) | Niti Aayog, Government of India | 0.12 | 0.11 |
| Healthcare access and quality index (HAQI) | Global Burden of Disease study | 0.50** | 0.35 |
| Vulnerability index (VI) | Population Council, India | 0.01 | 0.13 |
| p<0.05, ** p<0.01 |  |  |  |

**Figure 4:**
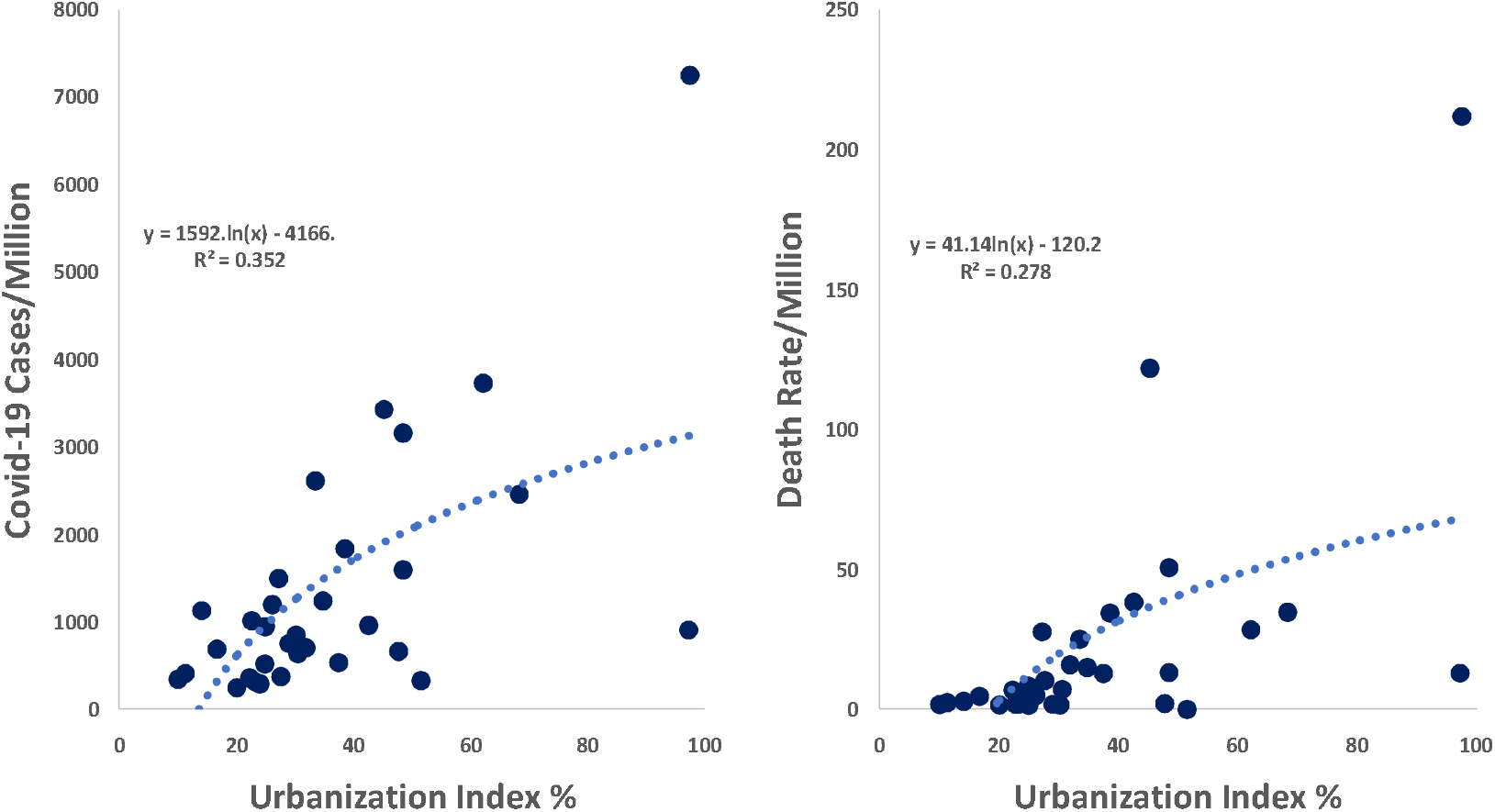
Association of Covid-19 cases and deaths/million with urbanization index (logarithmic regression) y = 41.14ln(x) – 120.2 R² = 0.278 s/M illion.

To evaluate monthly trends in association of cases and deaths/million with urbanization index we performed monthly analysis from April to July 2020 (Figure 5). Association of urbanization with cases/million for months of April, May, June and July, respectively, is similar (R^2^ = 0.38, 0.37, 0.38 and 0.29) while with deaths/million it is increasing (R^2^ = 0.10, 0.20, 0.21 and 0.28).

**Figure 5:**
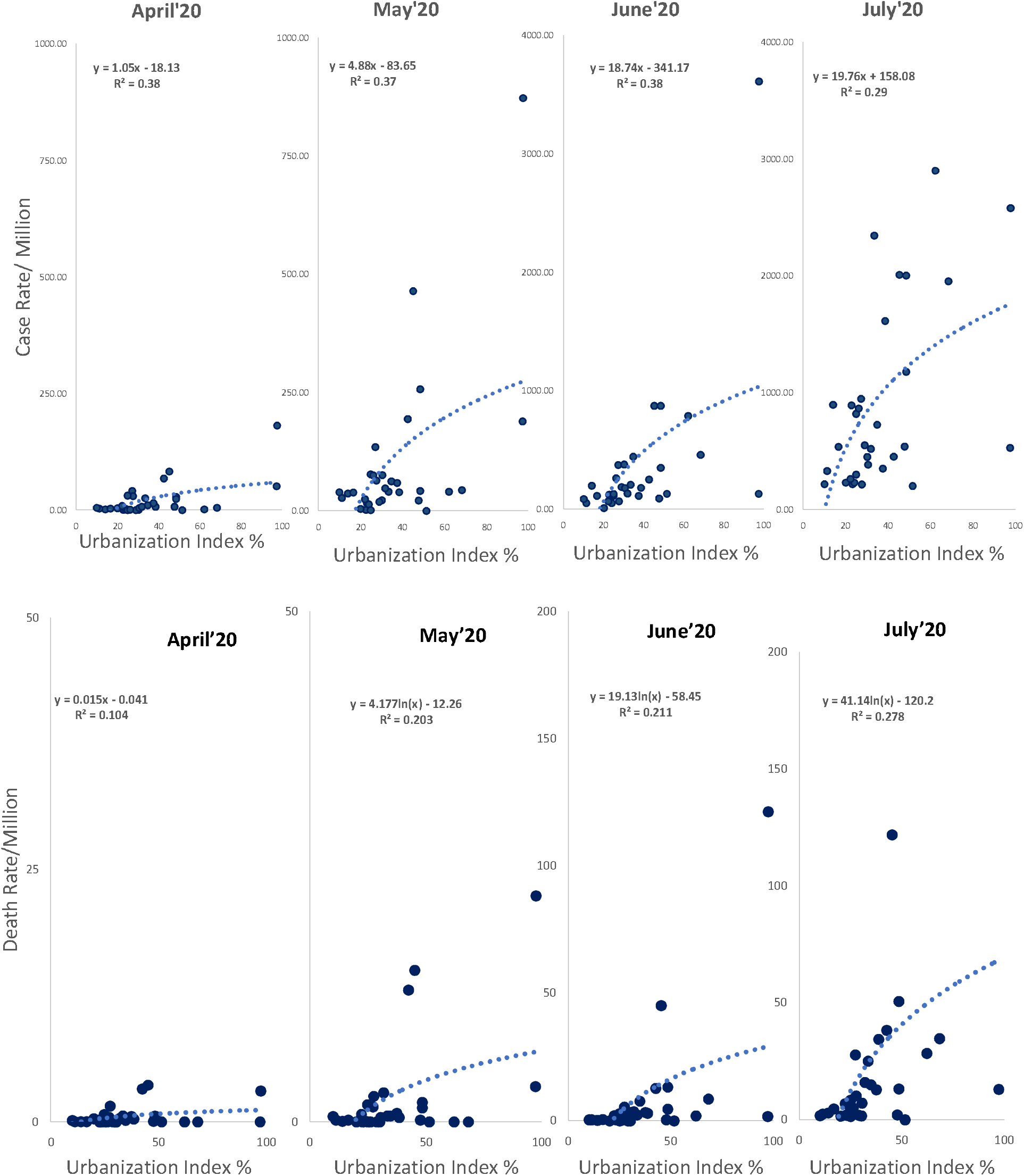
End-of-month logarithmic association of covid-19 cases/million and deaths/million with urbanization for April-July 2020 y = 1.05x – 18.13 R² = 0.38 n.

## DISCUSSION

This study shows rapidly increasing burden of covid-19 cases and deaths in terms of absolute numbers as well as proportion per million in all regions of India. The burden is significantly greater in more urbanized states and mortality from the disease is increasing in these states during the first six months of the epidemic.

Observational data from Europe and North America have not highlighted urban-rural difference in covid-19 burden.^5^ This could be due to the fact that most of these countries are highly urbanized (70− 90%). In countries with larger proportion of rural individuals, such as China, Brazil, Iran, Mexico and South Africa, reports have highlighted the predominant urban nature of the disease,^5^ similar to our study. Reports from China, Europe, UK and USA have also reported that urban low socioeconomic status individuals, especially the minorities, are at greater risk of disease and deaths from covid-19.^18,19,20^ The UK Office of National Statistics has reported that the age-standardized mortality rate from covid-19 in the most deprived areas was 55.1/100,000 compared to 25.3/100,000 in the least deprived areas.^21^ In USA, a study from New York reported that number of patients hospitalized/100,000 (moderate to severe disease) population was the highest in the borough of Bronx (634) and lowest in Manhattan (331), and number of deaths/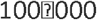 population was also the highest in Bronx (224) and lowest in Manhattan (122).^22^ The Bronx has the highest proportion of ethnic minorities, the most persons living in poverty and the lowest levels of educational attainment as compared to other boroughs of New York. Clinical registries from China, Europe and North America have highlighted a specific phenotype that is more prone to mortality from this infection- older age, male sex, hypertension, diabetes, obesity, concomitant cardiovascular disease and heart failure- especially in low socioeconomic status individuals.^19,20,23,24,25^ Because data on these variables is not available to us we cannot comment of association of these comorbidities with risk of developing or dying from covid-19 in India. However, epidemiological studies have consistently reported greater prevalence of elderly, obesity, hypertension, diabetes and cardiovascular diseases at the urban locations in India.^26,27,28,29^

In the present study we have not evaluated the patient-level socioeconomic or ethnic characteristics as such data are not available. However association of state-level development and healthcare related characteristics show insignificant correlation when adjusted for urbanization (Table 3). On the other hand, non-peer reviewed data from India has reported that more than two-thirds of the patients are concentrated in 13 Indian cities.^30^ Initial concentration of patients in low socioeconomic status localities within the urban locations has been reported.^9^ More studies regarding socioeconomic macro- and micro-level determinants of covid-19 infection are required from India as it has been predicted that covid-19 epidemic shall ultimately reside among the lower socioeconomic stratum in most countries.^12^

Public health strategies that have been successful in various high-income countries of Asia and Europe have focused on widespread testing for covid-19 virus using appropriate technologies, social (physical) distancing, quarantining of the cases using either hospital or home isolation and aggressive and rapid tracing of contacts and their isolation.^31^ Other interventions include protection of healthcare workers; monitoring hotspots; watching for imports; clear and honest communication; avoiding going back to ‘normal’; and investing in finding alternate strategies.^32^ Consideration for economic effects, social isolation, family relationships, health related behaviours, disruption to essential services, disruption of education, traffic, transport and green spaces, social disorder and psychosocial impacts is essential.^33^ Lockdown has led to both macro- micro-level adverse economic consequences in India and most developing countries.^34^ Development of preventive medicines and vaccines is a work in progress.^35^ Controlled initiation of population-wide (herd) immunity has been suggested as the only alternative for India,^36^ but if uncontrolled can lead to disastrous health consequences.

The essential urban nature of the disease and rapid spread of the disease in slums pose a challenge to control the epidemic in India and similar countries.^5^ Urban locations in India and slums are least prepared for the pandemic of covid-19 as most basic needs such as regular water supply, toilets, waste collection and adequate and secure housing are almost non-existent.^37^ Interventions that have been found useful in China and European countries may be impractical. Corburn et al suggest science-based policies for arresting course of the disease, improving general medical care, provision of economic, social and physical improvements, and focus on urban poor including migrants and slum-communities and certain microlevel strategies.^38^ Focus on using social and behavioural science techniques is essential to harness of benefits of such interventions.^39^ Focus on universal health coverage and creating resilient health infrastructure is crucial.^40,41^ Lessons from the covid-19 epidemic should guide policymakers to improve urban primary care and district hospitals.^42^

The present study shows that the covid-19 epidemic in India is still an urban phenomenon. It is probably in an intermediate stage of epidemiological transition.^9^ Non-pharmaceutical and multifocal policy level interventions are the most effective method to decrease the burden of this disease, although randomised trials are needed.^43^ A judicious strategy targeted to the urban population, especially the poor, with proper testing and containment, facilities for physical distancing and personal protection would be the most appropriate intervention.

## Data Availability

There are no data other than submitted in the manuscript.

http://www.covid19india.org

---

**Supplementary Table 1:** Demographic and social indices of various states

| State | Population 2020 | Urbanization index | Epidemiological transition index | Human development index | Socio-demographic index | Social Development Index | Health care availability index | Healthcare access & quality index | Vulnerability index |
| --- | --- | --- | --- | --- | --- | --- | --- | --- | --- |
| Data source |  | Census of India | Global Burden of Disease Study | Government of India | Global Burden of Disease Study | Government of India | Niti Aayog, Government of India | Global Burden of Disease Study | Population Council, India |
| Andhra Pradesh | 53903393 | 33.49 | 0.37 | 0.640 | 0.585 | 56.13 | 65.13 | 46.5 | 0.714 |
| Arunachal Pradesh | 1570458 | 22.67 | 0.55 | 0.660 | 0.589 | 55.24 | 46.07 | 44.3 | 0.029 |
| Assam | 35607039 | 14.08 | 0.62 | 0.614 | 0.562 | 48.53 | 48.85 | 34.0 | 0.257 |
| Bihar | 124799926 | 11.3 | 0.74 | 0.576 | 0.440 | 44.89 | 32.11 | 37.0 | 0.971 |
| Chandigarh | 1158473 | 97.25 | -- | 0.775 | -- | -- | 63.62 | 54.3 | 0.086 |
| Chhattisgarh | 29436231 | 23.24 | 0.6 | 0.613 | 0.558 | 56.69 | 53.36 | 37.4 | 0.314 |
| Delhi | 18710922 | 97.50 | 0.38 | 0.746 | 0.752 | 60.17 | 49.42 | 56.2 | 0.514 |
| Goa | 1586250 | 62.17 | 0.21 | 0.761 | 0.738 | 63.39 | 51.9 | 64.8 | 0.314 |
| Gujarat | 63872399 | 42.58 | 0.46 | 0.672 | 0.615 | 56.65 | 63.52 | 45.0 | 0.771 |
| Haryana | 28204692 | 34.79 | 0.4 | 0.708 | 0.655 | 57.37 | 53.51 | 45.0 | 0.400 |
| Himachal Pradesh | 7451955 | 10.04 | 0.3 | 0.720 | 0.669 | 65.39 | 62.41 | 51.7 | 0.057 |
| Jammu & Kashmir | 13606320 | 27.21 | 0.34 | 0.688 | 0.598 | 55.41 | 62.37 | 46.7 | 0.714 |
| Jharkhand | 38593948 | 24.05 | 0.69 | 0.599 | 0.523 | 47.8 | 51.33 | 37.4 | 0.914 |
| Karnataka | 67562686 | 38.57 | 0.34 | 0.682 | 0.614 | 59.72 | 61.14 | 46.6 | 0.571 |
| Kerala | 35699443 | 47.72 | 0.16 | 0.779 | 0.683 | 68.09 | 74.01 | 63.9 | 0.314 |
| Madhya Pradesh | 85358965 | 27.63 | 0.6 | 0.606 | 0.532 | 55.03 | 38.39 | 39.5 | 1.000 |
| Maharashtra | 123144223 | 45.23 | 0.33 | 0.696 | 0.666 | 57.88 | 63.99 | 49.8 | 0.829 |
| Manipur | 3091545 | 30.21 | 0.42 | 0.690 | 0.610 | 55.5 | 60.6 | 44.2 | 0.543 |
| Meghalaya | 3366710 | 20.08 | 0.64 | 0.656 | 0.585 | 53.51 | 55.95 | 39.6 | 0.286 |
| Mizoram | 1239244 | 51.51 | 0.53 | 0.705 | 0.630 | 62.89 | 74.97 | 48.9 | 0.143 |
| Nagaland | 2249695 | 28.97 | 0.47 | 0.679 | 0.661 | 56.76 | 38.51 | 46.1 | 0.657 |
| Orissa | 46356334 | 16.68 | 0.58 | 0.606 | 0.547 | 51.64 | 35.97 | 36.3 | 0.800 |
| Pondicherry | 1413542 | 68.31 | -- | 0.738 | -- | -- | 49.69 | -- | 0.200 |
| Punjab | 30141373 | 37.49 | 0.29 | 0.720 | 0.650 | 62.18 | 63.01 | 49.5 | 0.429 |
| Rajasthan | 81032689 | 24.89 | 0.66 | 0.629 | 0.530 | 52.31 | 43.1 | 40.7 | 0.686 |
| Sikkim | 690251 | 24.97 | 0.45 | 0.716 | 0.632 | 62.72 | 50.51 | 50.5 | 0.000 |
| Tamil Nadu | 77841267 | 48.45 | 0.26 | 0.708 | 0.648 | 65.34 | 60.41 | 51.2 | 0.571 |
| Telangana | 39362732 | 48.45 | 0.38 | 0.669 | 0.609 | -- | 59 | 48.5 | 0.943 |
| Tripura | 4169794 | 26.18 | 0.45 | 0.663 | 0.573 | 53.22 | 46.38 | 42.3 | 0.629 |
| Uttar Pradesh | 237882725 | 22.28 | 0.68 | 0.596 | 0.508 | 50.96 | 28.61 | 34.9 | 0.886 |
| Uttarakhand | 11250858 | 30.55 | 0.46 | 0.684 | 0.639 | 64.23 | 40.2 | 43.2 | 0.429 |
| West Bengal | 99609303 | 31.87 | 0.33 | 0.641 | 0.580 | 54.37 | 57.17 | 47.1 | 0.829 |

